# Future-Information Leakage in 30-Day Readmission Prediction: An Empirical Audit and a Calibrated Electronic Health Record Model in Medicare-Insured Adults

**DOI:** 10.64898/2026.11.13.26368026

**Authors:** Thiago Bandeira, Armando Gonzalez, Christian Poellabauer, Ananda Mohan Mondal

## Abstract

**Objective:** To develop and internally validate a calibrated discharge model of 30-day within-system readmission in Medicare-insured adults, and to quantify how a future-information leak and outcome artifacts distorted performance, timing, and subgroup conclusions.

**Materials and Methods:** Retrospective study of 236,906 Medicare-insured MIMIC-IV v3.1 admissions (87,750 patients; prevalence 21.4%). Predictors were audited for discharge-time availability; history features were tested for future information and recomputed from raw timestamps. A consensus gradient-boosted (XGBoost) model was selected inside patient-grouped folds; CIs used patient-cluster bootstrap.

**Results:** A feature documented as a prior 180-day length-of-stay trend used each patient’s whole record, including later admissions; removing it and two derived terms lowered test area under the receiver operating characteristic curve (AUROC) from 0.774 to 0.706. The corrected procedure reached an out-of-fold AUROC of 0.7057 (95% CI 0.7016 to 0.7102; primary); the 44-feature model scored 0.7056 on the historically exposed test partition (calibration slope 0.99), above reconstructed LACE and HOSPITAL (both P<.001). A day-1 discrimination peak largely reflected 2,368 probable administrative continuations within 1 hour of discharge; merging these into the preceding stay gave test AUROC 0.698 (0.702 after refitting). The White minus Black AUROC difference was 0.003 (95% CI −0.018 to 0.024); specificity was lower for Black patients (0.61 vs 0.69).

**Discussion:** Documentation audits and patient-grouped validation missed a leak that inflated discrimination and created spurious subgroup and calendar-period differences; simple empirical checks found it.

**Conclusion:** Empirical leakage checks should accompany availability audits. The corrected model is calibrated, moderately discriminating, and needs external validation before clinical use.

## Background and Significance

Unplanned 30-day readmissions burden the United States health system, and the Hospital Readmissions Reduction Program (HRRP) of the Centers for Medicare & Medicaid Services has tied reimbursement to risk-adjusted readmission performance since 2013 [1,2]. Risk estimates available at discharge could target transitional care if the model can be trusted at deployment. Timing and equity sharpen the stakes: early and late readmissions differ in causes [3,4], and unequal model performance across groups can lead to unequal care [5,6].

Bedside indices remain the default. The LACE index [7] and the HOSPITAL score [8] reach AUROCs of about 0.68 to 0.77 in derivation and validation studies [7–10]; a systematic review found that most readmission risk models perform poorly [11], and reviews note recurring weaknesses: leakage-prone splits, absence of calibration assessment, and unexamined subgroup performance [11–13]. Machine learning on EHR data can improve on HOSPITAL-type scores [14], and gradient-boosted trees remain strong learners on tabular clinical data [15–18]. Clinical use also requires calibrated probabilities [19], interpretable explanations [20], same-cohort comparison with the scores hospitals use, fairness auditing [5,21–23], and reporting aligned with TRIPOD+AI [24] and PROBAST+AI [25].

Data leakage, the use of information during development that would not be available at prediction time, is a leading cause of overoptimistic and irreproducible results in machine-learning science [26]. EHR prediction is especially exposed: billing codes finalized after discharge, preprocessing fit on evaluation data, splits that ignore patient clustering, and engineered features whose time windows are not what their names suggest [25,27]. Roadmaps call for patient-level partitioning, fold-local preprocessing, and verified predictor availability [24,25,28], but verification usually means reading documentation rather than testing the data.

On the Medical Information Mart for Intensive Care (MIMIC-IV) [29], published 30-day readmission models report AUROCs of 0.70 to 0.79 [30–32], and ClinicalBERT about 0.714 on MIMIC-III notes [33]; none combines leakage-safe validation with empirical leakage tests, calibration, same-cohort clinical-score comparison, timing analysis, and a subgroup audit, which this study does in the Medicare population.

### Objective

Five research questions (RQs) organize the study. RQ1: Which discharge-available EHR features contribute most to prediction of 30-day readmission in Medicare-insured adults? RQ2: Can a leakage-safe, calibrated machine-learning model improve prediction relative to established clinical scores? RQ3: Can global Shapley additive explanations (SHAP) provide clinically interpretable insight into predicted risk? RQ4: How does performance vary across the post-discharge time window and across demographic subgroups? RQ5: How did a future-information leak and administrative outcome artifacts distort performance, timing, and subgroup conclusions, and can simple empirical checks detect them?

## Materials and Methods

### Study Design and Data Source

We conducted a retrospective prediction-model development and internal-validation study reported in line with TRIPOD+AI [24] (checklist: Supplementary Appendix 5; PROBAST+AI self-assessment [25]: Supplementary Appendix 6). The data source is MIMIC-IV v3.1 [29,34], a deidentified EHR database of 546,028 hospitalizations between 2008 and 2022 at Beth Israel Deaconess Medical Center (Boston, MA); dates are shifted per patient with within-patient intervals preserved. Clinical notes and imaging were not used.

### Cohort and Outcome Definition

The cohort comprised admissions with Medicare insurance (n=244,576). Because the model scores patients at alive discharge, 7,670 admissions ending in index in-hospital death were excluded, leaving 236,906 admissions from 87,750 patients. Medicare insurance does not imply age 65 or older, and fee-for-service cannot be distinguished from Medicare Advantage, so this is not an HRRP regulatory cohort. The outcome was the first same-system readmission for any cause within 0 to 30 days after index discharge, identified from all subsequent hospitalizations irrespective of insurance. Next admissions overlapping the index discharge by up to 2 days (n=650) were treated as transfers. Every index admission is eligible, so patients contribute multiple admissions; all inferences account for this clustering. Planned readmissions could not be reliably identified; sensitivity analyses vary the outcome definition (Supplementary Methods S1).

### Partitions and Validation Design

Admissions were split 80/20 at the patient level into development and test data, with 10% of the development portion assigned, again patient-grouped, to a validation partition used for the operating threshold and for early stopping of the survival model. After exclusions: training 170,677 admissions (63,155 patients, 36,665 events), validation 18,557, and test 47,672 (17,553 patients, 10,133 events). Earlier development phases scored model variants on the test partition repeatedly, so it is a historically exposed internal partition. Three evidence tiers are reported: (1) primary, out-of-fold performance of the complete selection procedure under 5 outer patient-grouped folds on the development partitions; (2) secondary, grouped cross-validation of the fixed consensus set (optimistic, because the set aggregates selection across folds); and (3) tertiary, one evaluation of the final model on test, used for detailed metrics, subgroups, baselines, and timing. No test outcome entered this version’s selection, encoding, or threshold.

### Predictor Audit, Leakage Testing, and Feature Selection

All 207 candidate predictors were audited for provenance, measurement window, missingness, and documented availability at discharge (Supplementary Appendix 1). Sixty-three billing-derived features (diagnosis-related groups, International Classification of Diseases (ICD) codes, and their derivatives), one race-derived encoding, and one precomputed encoding of discharge destination were excluded. Because documentation can be wrong, we added three empirical checks for information from later encounters. First, among first observed admissions (no earlier admission in MIMIC-IV), a feature built only from prior stays carries no information, so a feature fixed at one value for every patient without a later admission, but not for others, encodes the future. Second, a feature computed once per patient over the whole record is identical across that patient’s admissions. Third, every history feature of the final model was recomputed from raw admission timestamps; laboratory, chart, medication-administration, and prescription features were rebuilt from raw records that both occurred and were documented (stored or entered) on or before discharge; two exact duplicate features were dropped; and outpatient and other index-stay features were checked against raw tables (Supplementary Methods S3). These checks target one class of leakage (information from later encounters) and cannot certify that every predictor is leak-free.

Feature selection used staged recursive feature elimination (RFE) with an XGBoost estimator inside 5 outer patient-grouped folds on development data only. Within each outer-training fold, 3 grouped inner folds drove elimination along a fixed grid; categorical variables were target-encoded from inner-training patients only; and a parsimony rule, fixed before this analysis version, chose the smallest feature count within 0.002 inner-validation AUROC of the best. Features selected in at least 3 of 5 folds formed the consensus set. The source feature table had imputed 16 continuous measurements with medians computed over all partitions, including test, and had zero-filled 5 others; all were rebuilt from raw tables with missing values left missing for the tree learner, counts set to zero when no event occurred, and time since last discharge left missing for first observed admissions (Supplementary Methods S4, Table S3).

### Model and Comparators

The final model is a single XGBoost classifier [16] on the consensus set (600 trees without early stopping, learning rate 0.05, maximum depth 5, subsample and column subsample 0.9, fixed seed), refit on the training partition. Its operating threshold was the validation-partition Youden point (0.207), development-derived and applied unchanged to test. Comparators fitted per outer fold on identical data were the full 137-feature eligible pool, the clean subset of an earlier development feature set, the earlier 66-feature set (containing billing features and leaked variables; historical reference), and regularized logistic regression.

### Clinical-Score Baselines

LACE [7] and HOSPITAL [8] were reconstructed for every admission, HOSPITAL with its original 12-month prior-admission window and LACE in adapted form, because emergency department visits that did not lead to admission are absent from the MIMIC-IV core tables (Supplementary Appendix 2). Both were compared with the out-of-fold predictions on the development partitions and with the final model on test. Two components use coded data from the index stay (LACE comorbidity from ICD diagnoses; the HOSPITAL procedure item from ICD procedures), information our model was not allowed to use. Because the integer scores were derived for other outcomes, we also refit their components by logistic regression on the training partition, with and without the coded components.

### Leakage Ablation

To quantify how common pipeline choices inflate apparent performance, we compared the cross-validated (apparent) AUROC on development data with the held-out test AUROC of the model each choice would deploy.

### Time-to-Event Analyses

Time to readmission used a competing-risks event process (readmission; death before readmission from hospital and Massachusetts registry records; administrative censoring at day 30) with Aalen-Johansen cumulative incidence and a cause-specific XGBoost accelerated failure time (AFT) model [35]. Evaluation used Harrell C [36] and cumulative/dynamic AUROC on days 1 to 29 [37,38], with patients who died before the evaluation day kept among the controls [38]; with no censoring before day 30 the estimates are empirical, with patient-cluster bootstrap bands. Three landmark models covered days 1 to 7, 8 to 14, and 15 to 30.

### Statistical Analysis

All confidence intervals (CIs) and P values use patient-level cluster bootstrap percentile resampling (1,000 resamples for the test panel; 5,000 for contrasts with reported P values; two-sided empirical P values with plus-one correction). Model-to-model contrasts are paired; the White vs Black contrast resamples patients jointly, so a patient recorded in both groups receives the same multiplicity in both. Formal testing was limited to three contrasts carried over unchanged from the analysis plan of the earlier, leaked version (vs LACE, vs HOSPITAL, and White vs Black); conclusions are unchanged under Bonferroni adjustment for three tests. All other comparisons, including component refits and subgroup differences, are descriptive and reported with CIs only. Calibration used the Brier score, expected calibration error (ECE) over 10 equal-width bins, the calibration slope, and calibration-in-the-large (intercept with slope fixed at 1) [19]. Decision-curve analysis [39] covered thresholds 0.05 to 0.40. Analyses used Python 3.11.14 with XGBoost 3.2.0, scikit-learn 1.7.2, pandas 2.3.3, and NumPy 1.26.4 (seed 42).

### Subgroup Audit

Performance was audited on the test partition by age band, sex, race, and number of prior admissions (0, 1 to 2, 3 to 9, 10 or more), with race strings consolidated by substring mapping (Supplementary Appendix 3). Race is not a model input; auditing by race is deliberate because omitting an attribute does not guarantee equal performance [5,21,22].

### Ethical Considerations

The institutional review board of Beth Israel Deaconess Medical Center granted a waiver of informed consent for MIMIC-IV and approved its sharing [29]. The first author accessed the data under the PhysioNet Credentialed Health Data Use Agreement after required training; co-authors used only aggregate results, and no row-level data are redistributed. This secondary analysis of deidentified data did not require institutional review board review at Florida International University. No protocol was prepared, the study was not registered, and patients and the public were not involved.

## Results

### Cohort

Figure 1 shows the cohort flow. Of 244,576 Medicare admissions, 7,670 ended in index in-hospital death, leaving 236,906 admissions from 87,750 patients; 50,793 (21.4%) were followed by a within-system readmission within 30 days. Partitions were similar in case mix (Table 1).

**Figure 1.**
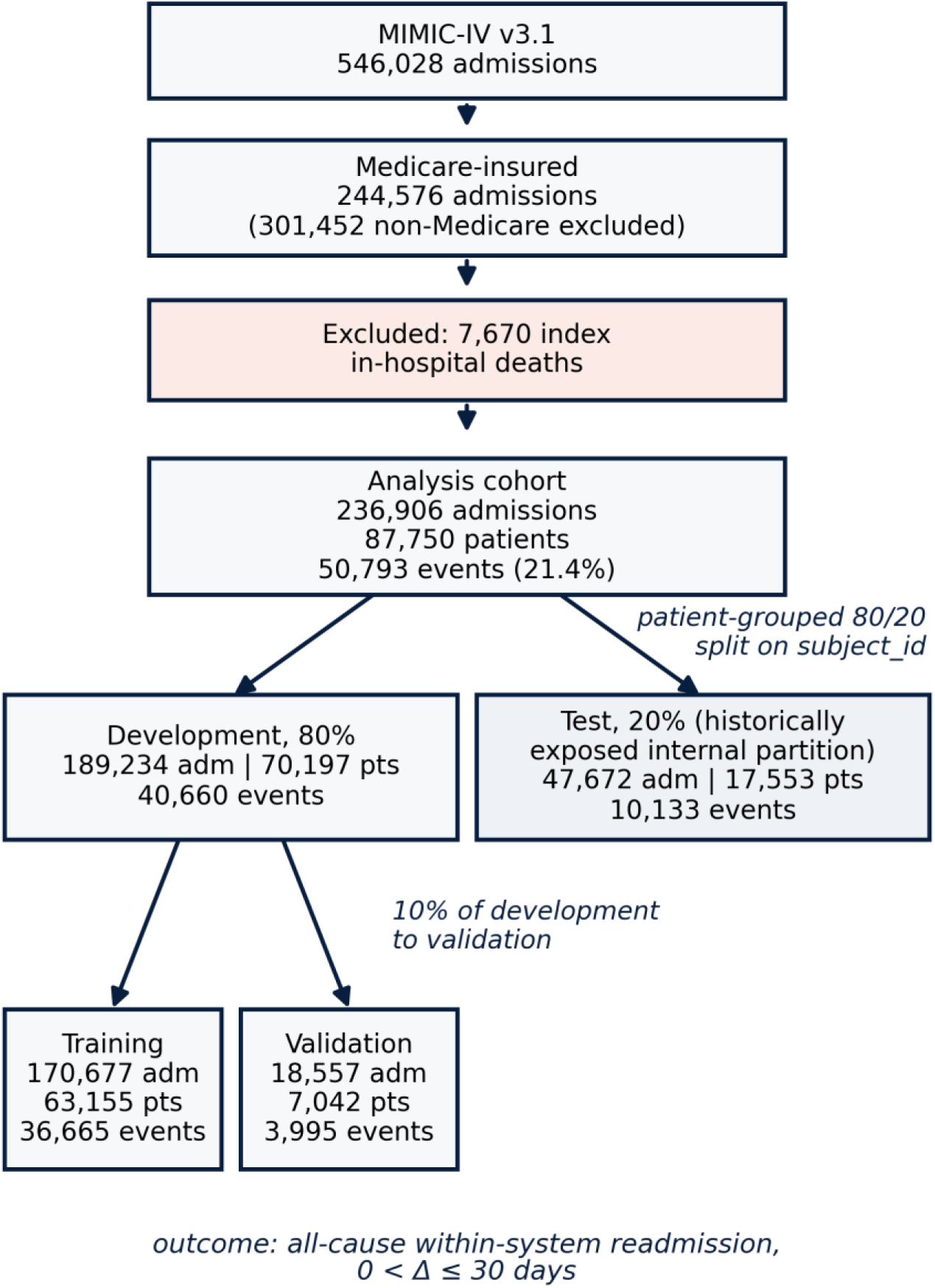
Participant flow with patient-grouped partitions; feature selection ran inside 5 patient-grouped folds drawn from the development partitions. Alt text: Flow diagram. 546,028 MIMIC-IV hospitalizations; 244,576 with Medicare insurance; 7,670 index in-hospital deaths excluded; 236,906 admissions from 87,750 patients analyzed, split by patient into training, validation, and test partitions.

**Table 1.** Cohort characteristics by partition. Values are n (%) or median (interquartile range, IQR). Race groups follow the consolidation in Supplementary Appendix 3. Prior admissions count earlier admissions of any insurance in MIMIC-IV. Per-predictor missingness appears in Supplementary Appendix 1.

| **Characteristic** | **Training** | **Validation** | **Test** | **All** |
|---|---|---|---|---|
| Admissions | 170,677 | 18,557 | 47,672 | 236,906 |
| Patients | 63,155 | 7,042 | 17,553 | 87,750 |
| 30-day readmission (events) | 36,665 (21.5%) | 3,995 (21.5%) | 10,133 (21.3%) | 50,793 (21.4%) |
| Age, years, median (IQR) | 74 (66 to 82) | 74 (66 to 82) | 74 (66 to 82) | 74 (66 to 82) |
| Age under 65 | 35,916 (21.0%) | 3,916 (21.1%) | 10,130 (21.2%) | 49,962 (21.1%) |
| Female | 88,001 (51.6%) | 9,730 (52.4%) | 24,465 (51.3%) | 122,196 (51.6%) |
| White | 125,745 (73.7%) | 13,739 (74.0%) | 35,059 (73.5%) | 174,543 (73.7%) |
| Black | 24,340 (14.3%) | 2,515 (13.6%) | 6,715 (14.1%) | 33,570 (14.2%) |
| Hispanic or Latino | 6,269 (3.7%) | 696 (3.8%) | 1,993 (4.2%) | 8,958 (3.8%) |
| Asian | 3,691 (2.2%) | 399 (2.2%) | 1,101 (2.3%) | 5,191 (2.2%) |
| Other or unknown race | 10,632 (6.2%) | 1,208 (6.5%) | 2,804 (5.9%) | 14,644 (6.2%) |
| Elective admission | 4,538 (2.7%) | 438 (2.4%) | 1,224 (2.6%) | 6,200 (2.6%) |
| Any ICU stay | 28,993 (17.0%) | 3,189 (17.2%) | 8,202 (17.2%) | 40,384 (17.0%) |
| Length of stay, days, median (IQR) | 3.0 (1.0 to 6.0) | 3.0 (1.0 to 6.0) | 3.0 (1.0 to 6.0) | 3.0 (1.0 to 6.0) |
| Prior admissions, median (IQR) | 1 (0 to 4) | 1 (0 to 4) | 1 (0 to 5) | 1 (0 to 4) |
| No prior admission | 56,449 (33.1%) | 6,331 (34.1%) | 15,639 (32.8%) | 78,419 (33.1%) |
| Admission in prior 180 days | 69,584 (40.8%) | 7,453 (40.2%) | 19,438 (40.8%) | 96,475 (40.7%) |
| No laboratory results recorded | 20,080 (11.8%) | 2,340 (12.6%) | 5,836 (12.2%) | 28,256 (11.9%) |

### Future-Information Leakage

The empirical test flagged one documented prior-utilization feature, a 180-day length-of-stay trend that had been the strongest predictor of an earlier version of this model. Among 78,419 first observed admissions, the trend was nonzero for 44.2%; every such patient had a later admission, and no patient without one had a nonzero value (30-day readmission 29.8% vs 3.1%). It was also identical across all admissions for each of the 45,154 multi-admission patients: we computed it once per patient over the whole record. It and two interaction terms were removed. No other candidate showed the first-admission signature (sex, recorded once per patient, is constant for a legitimate reason), and every history feature of the final model was reproduced from earlier admissions only (99.7% to 100% of admissions). Removing the three features lowered the out-of-fold AUROC from 0.7748 to 0.7057 and test AUROC from 0.7738 to 0.7056. A correctly built trend, using only stays completed before the index admission, added no discrimination to the final features (development cross-validation AUROC 0.7056 vs 0.7057; Supplementary Methods S3), so the inflation came from the leak itself. A second defect was found by comparing stored columns with raw tables: 16 continuous measurements had been imputed with medians computed over all partitions and 5 zero-filled, so values were present for admissions with no measurement (Supplementary Methods S4); the table was rebuilt with missing values preserved and a discharge cutoff enforced on every laboratory and medication count, and all results below use it and the corrected pool of 137 features.

In the ablation (Table 2), adding the trend features raised apparent and held-out AUROC alike (by 0.071 and 0.069), because the future information was present in the test data too: internal validation cannot detect this kind of leak. Combining all choices produced apparent AUROC 0.806 against 0.768 held out.

**Table 2.** Leakage ablation. Apparent AUROC is the 5-fold cross-validated estimate on development data under each pipeline choice; held-out AUROC is that of the model the choice would deploy, trained on the training partition and scored on the test partition. Optimism is apparent minus held-out. CV: cross-validation.

| **Pipeline choice** | **Features** | **Apparent CV AUROC** | **Held-out test AUROC** | **Optimism** |
|---|---|---|---|---|
| Leakage-safe reference | 44 | 0.7057 | 0.7056 | +0.0001 |
| Admission-level split | 44 | 0.7056 | 0.7056 | +0.0000 |
| Encoding fit on all development data | 44 | 0.7059 | 0.7056 | +0.0003 |
| Add future-information trend features | 47 | 0.7771 | 0.7745 | +0.0026 |
| Add legacy precomputed encodings | 48 | 0.7131 | 0.7114 | +0.0018 |
| Add billing-derived features | 104 | 0.7071 | 0.7090 | −0.0019 |
| All of the above combined | 111 | 0.8063 | 0.7680 | +0.0383 |

### Feature Selection and Model Comparison

The parsimony rule selected 45, 38, 55, 45, 45 features across the outer folds (mean pairwise Jaccard 0.771; 34 selected in all 5; Figure 2), and the consensus rule yielded 44 features. Out of fold, the selection procedure achieved AUROC 0.7057 (95% CI 0.7016 to 0.7102), equal to the full 137-feature pool (0.7064; paired difference 0.0007, 95% CI −0.0001 to 0.0014) and above regularized logistic regression (0.6860); the earlier 66-feature set, which contains the leaked trend, scored 0.7637.

**Figure 2.**
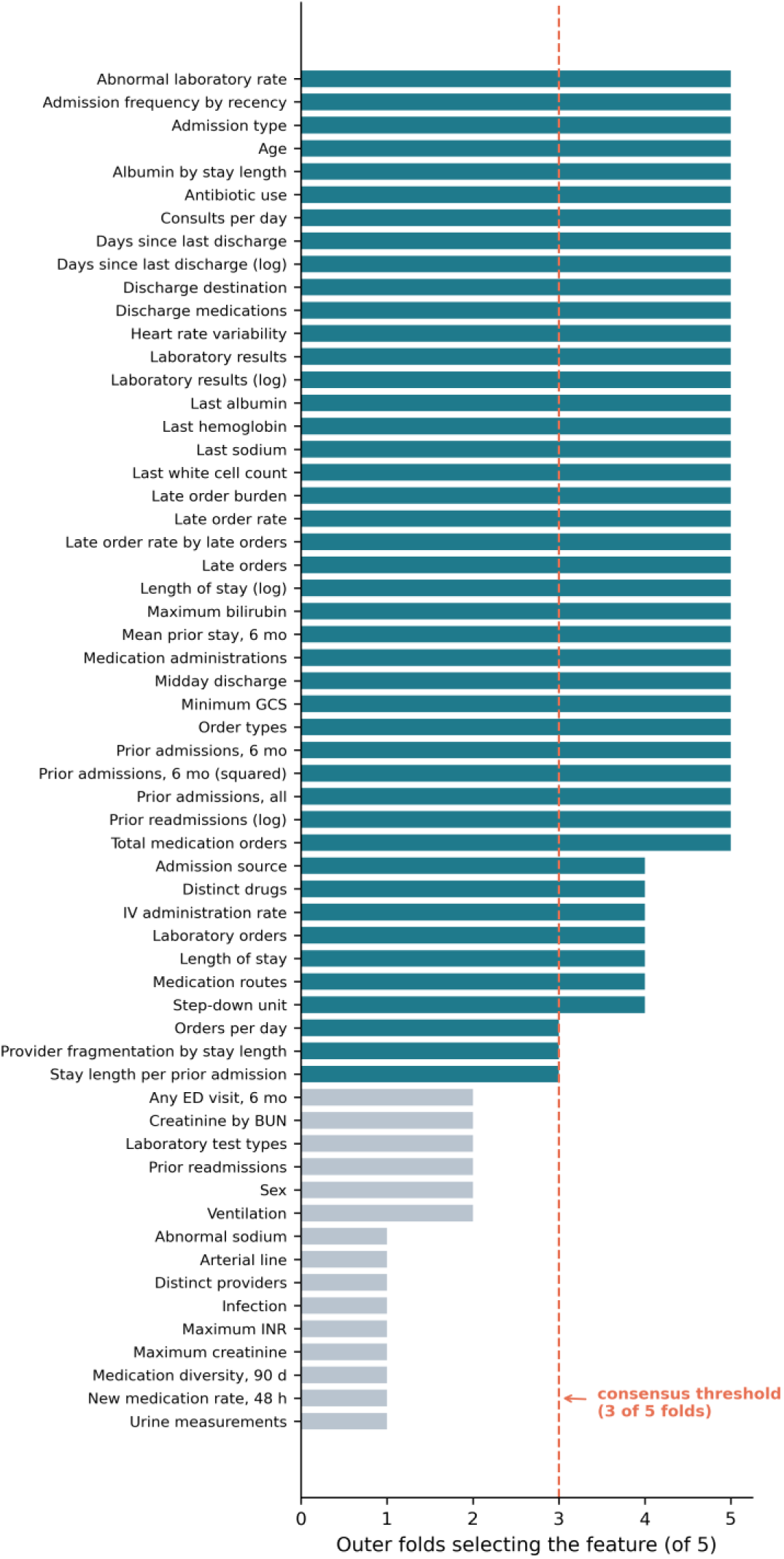
Selection frequency across the 5 outer folds for all 59 features selected at least once; teal bars form the consensus set, and the dashed line marks the consensus threshold (3 of 5 folds). Alt text: Horizontal bar chart of the number of outer folds (0 to 5) selecting each of 59 features; the 44 consensus features, selected in 3 or more folds, are teal and the others gray; a dashed line marks 3 folds.

### Discrimination and Calibration

Out of fold, the selection procedure’s predictions were also calibrated (slope 1.00, Brier 0.1502, ECE 0.002) with average precision 0.430; the top decile captured 25% of events at a positive predictive value of 0.53. Development cross-validation of the fixed consensus model gave AUROC 0.7057 (fold SD 0.0020). Every result below that is not labeled out of fold comes from the exposed test partition (tertiary evidence). On it, the model reached AUROC 0.7056 (95% CI 0.6964 to 0.7145) and average precision 0.429 against a 21.3% prevalence. Calibration was close to ideal: slope 0.99 (95% CI 0.96 to 1.02), calibration-in-the-large −0.01 (95% CI −0.03 to 0.01; mean predicted 21.4% vs observed 21.3%), ECE 0.003, and Brier 0.1489 (Figure 3). At the threshold of 0.207, sensitivity was 0.624, specificity 0.674, positive predictive value 0.341, and negative predictive value 0.869.

**Figure 3.**
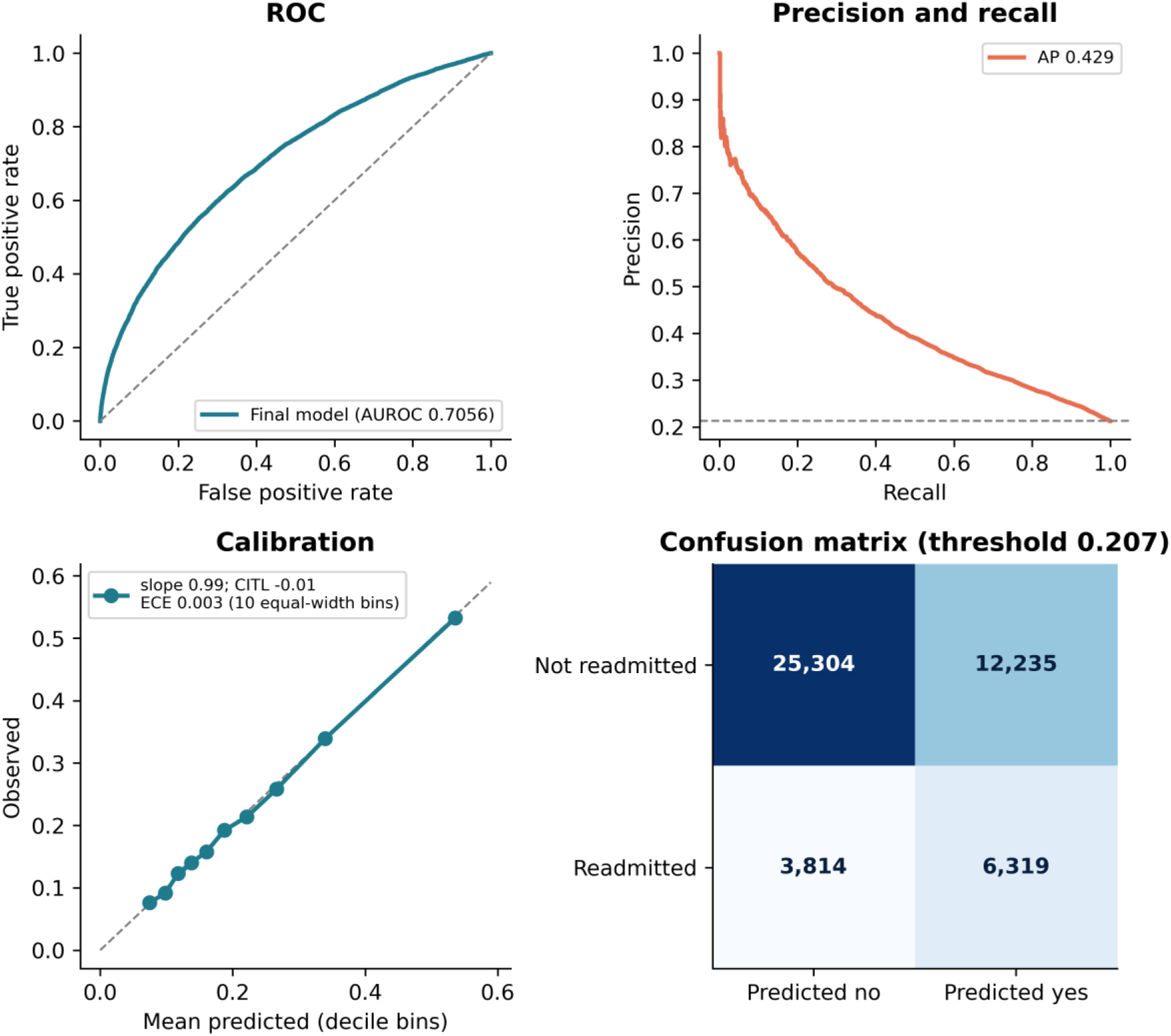
Receiver operating characteristic, precision and recall, calibration, and confusion-matrix panels for the final model on the test partition. Calibration points are decile bins of predicted risk; ECE uses 10 equal-width bins; CITL: calibration-in-the-large. Alt text: Four panels for the test partition: a receiver operating characteristic curve with AUROC 0.706; a precision-recall curve with average precision 0.429; a calibration plot whose decile points lie close to the diagonal; and a two-by-two confusion matrix at threshold 0.207.

### Sensitivity Analyses

Counting the transfer band as readmissions (refit AUROC 0.7101), excluding next admissions typed ELECTIVE (refit 0.7000), restricting to nonelective index admissions (final model 0.7073), and excluding admissions reaching the 2022 end of data (final model 0.7059) left discrimination essentially unchanged. Under the 1-hour episode rule (Timing of Readmission; flow in Supplementary Table S4), the frozen final model scored 0.6977 (95% CI 0.6880 to 0.7054) and a model refit to this outcome reached 0.7019. AUROC was stable across calendar-period bands (0.693 to 0.706; Supplementary Table S1; before the correction, 0.744 to 0.856).

### Same-Cohort Clinical Baselines

Out of fold on the development partitions, the selection procedure exceeded adapted LACE (0.6055) by 0.1002 (95% CI 0.0961 to 0.1041) and HOSPITAL (0.6424) by 0.0634 (95% CI 0.0607 to 0.0662; paired, descriptive), and its top decile captured 25% of events against 16% for LACE and 20% for HOSPITAL. On test, the final model outperformed adapted LACE (0.6050) by 0.1006 (95% CI 0.0916 to 0.1098; P<.001) and HOSPITAL (0.6468) by 0.0587 (95% CI 0.0530 to 0.0644; P<.001). Refitting the score components to this cohort raised LACE to 0.6400 and HOSPITAL to 0.6592; combining the components of both without the coded items gave 0.6647, still below the model by 0.0409 (95% CI 0.0361 to 0.0456; descriptive) (Table 3). These margins compare a model built for this outcome with scores derived for other outcomes, two of whose items use coded data our model excluded; statistical significance does not establish a clinically meaningful improvement.

**Table 3.** Comparison with clinical scores, a linear model, and published MIMIC-IV readmission models. Published values come from different cohorts and outcome definitions and are reference points, not head-to-head results. Rows for this study are in bold.

| Comparator | Data | AUROC | Note |
|---|---|---|---|
| Adapted LACE, integer score [7] | Development, out of fold | 0.6055 | ED visits without admission not captured |
| HOSPITAL, integer score [8] | Development, out of fold | 0.6424 | 12-month window |
| Adapted LACE, integer score [7] | This test partition | 0.6050 | Reconstructed; uses coded comorbidity |
| LACE, components refit | This test partition | 0.6400 | Logistic refit on training data |
| HOSPITAL, integer score [8] | This test partition | 0.6468 | 12-month window; uses coded procedures |
| HOSPITAL, components refit | This test partition | 0.6592 | Logistic refit on training data |
| LACE and HOSPITAL, refit without coded items | This test partition | 0.6647 | Discharge-available components only |
| Logistic regression, eligible pool | Development, out of fold | 0.6860 | 137 features |
| **This study, selection procedure** | **Development, out of fold** | **0.7057** | **Primary evidence** |
| **This study, fixed 44-feature model** | **Test partition** | **0.7056** | **Tertiary evidence** |
| Adisa 2026 [32] | MIMIC-IV, all adults | 0.696 | Different cohort and outcome |
| Almeida 2025 [31] | MIMIC-IV, all adults | 0.727 | Discharge notes; different cohort |
| Tang 2023 [30] | MIMIC-IV, radiograph subset | 0.791 | Multimodal; imaging-selected |

### Timing of Readmission

The test partition contributed 10,117 readmissions and 1,170 deaths before readmission (30-day cumulative incidence 21.2% and 2.5%). The AFT model reached Harrell C 0.6757. Daily AUROC peaked on day 1 (0.836, 95% CI 0.815 to 0.854) and fell to about 0.69 by day 7 (Figure 4). The peak was an artifact: 2,368 readmissions (4.7% of events) began within 1 hour of discharge, a rate of 2,368 per hour against 41 to 116 per hour at every later interval. Most appeared administrative (51% internal psychiatric transfers, 14% arrivals from skilled nursing, 40% with an emergency department registration before the index discharge), which is consistent with, but does not confirm, continuation; because the 1-hour window was derived from the data, the primary outcome was kept and the rule is a sensitivity analysis (Supplementary Methods S1, Table S4). It merges admissions beginning within 1 hour of a discharge into one stay, starts the 30-day window when that stay ends, excludes stays ending in death, merges continuation rows rather than scoring them as separate index admissions, and keeps the predictions made at the first discharge, a different prediction setting from the primary analysis. Under this rule the day-1 AUROC fell to 0.683 (95% CI 0.649 to 0.719), still slightly above later days but with an interval that overlaps them; discrimination was lowest in the first week (minimum 0.636) and rose to 0.673 by day 29. Landmark AUROCs under this rule were 0.662, 0.687, and 0.711 for days 1 to 7, 8 to 14, and 15 to 30.

**Figure 4.**
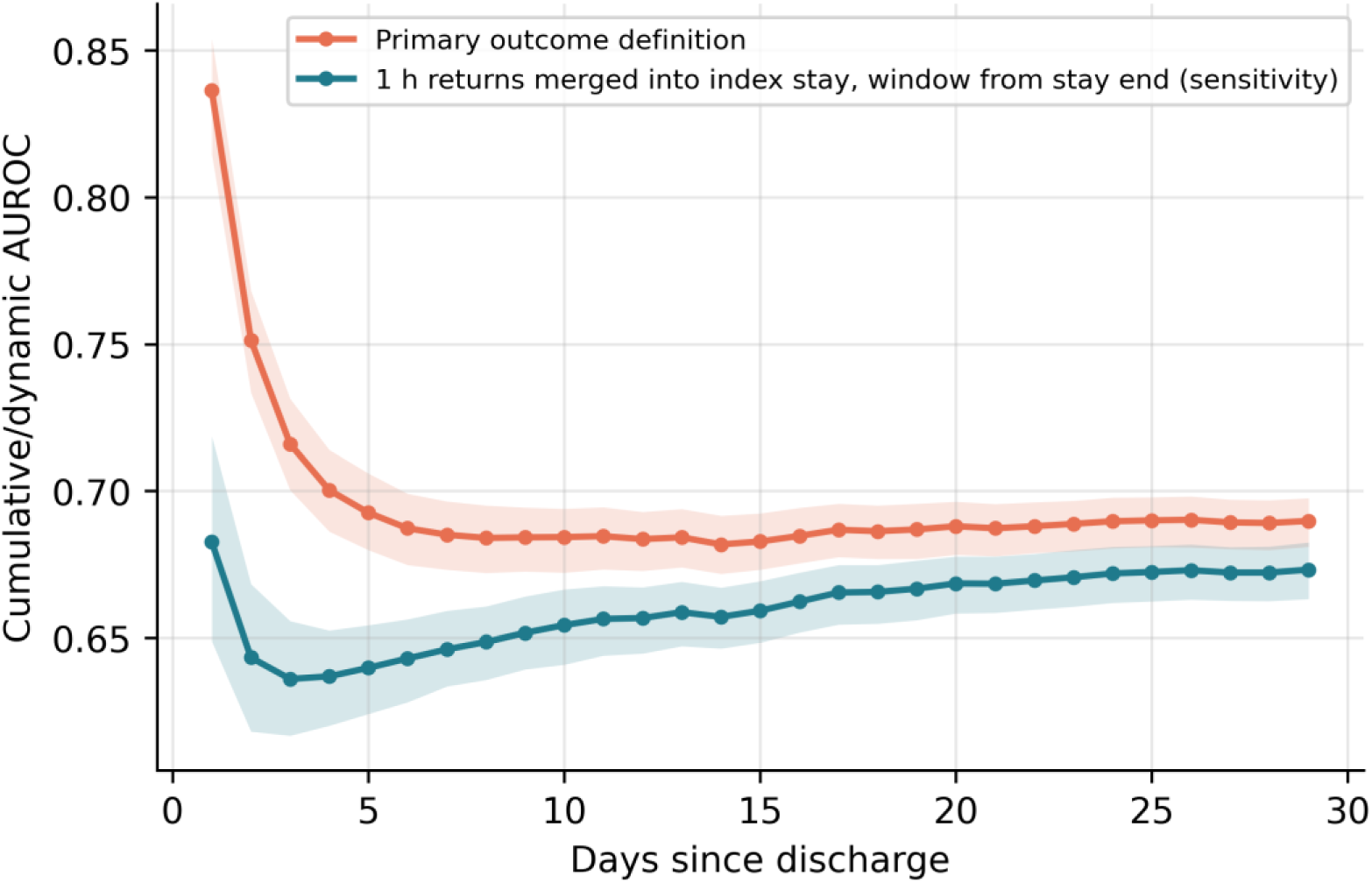
Cumulative/dynamic AUROC of the AFT risk score by day since discharge, with deaths before each day kept among the controls; shaded bands are 95% patient-cluster bootstrap intervals. The primary outcome definition counts returns beginning within 1 hour of discharge as readmissions; the second curve treats them as part of the index stay, with the 30-day window starting when that stay ends (sensitivity analysis). Alt text: Line chart of AUROC by day 1 to 29 with shaded 95% bands for two outcome definitions. The primary curve starts at 0.84 on day 1 and falls to about 0.69; the sensitivity curve starts at 0.68, dips to 0.64 in the first week, and rises to 0.67 by day 29.

### Subgroup Performance

No difference in discrimination between Black and White patients was detected (Table 4; AUROC 0.6948 vs 0.6976; difference 0.0029, 95% CI −0.0184 to 0.0237; P=.78), but the interval does not exclude differences of up to about 0.02, so this is not evidence of equivalence. Before the leak was removed, this difference had been 0.034 (P<.001), because the leaked trend was informative mainly for patients with few prior admissions, who are more often White (median prior admissions 1 vs 3). Discrimination was lower in older patients (0.6823 for ages 75 to 84 and 0.6716 for 85 or older, vs 0.7208 for 65 to 74) and rose with prior utilization, from 0.6573 (95% CI 0.6439 to 0.6695) for first admissions, 33% of the test partition, to 0.6987 for patients with 10 or more prior admissions, who formed 46% of the highest-risk decile (Figure 5, Supplementary Table S2). Within strata the White minus Black difference remained small (standardized 0.001, 95% CI −0.017 to 0.022). At the shared threshold, specificity was 0.610 for Black and 0.685 for White patients (difference 0.075, 95% CI 0.048 to 0.102; descriptive), so nonreadmitted Black patients were flagged more often. Mean predicted risk matched observed risk for Black and White patients (observed to expected 1.00 and 1.00), but the Black calibration slope was below 1 (0.921, 95% CI 0.839 to 0.999), indicating slightly too extreme risks, and risk was overestimated for the heterogeneous other or unknown group (observed to expected 0.78).

**Table 4.**
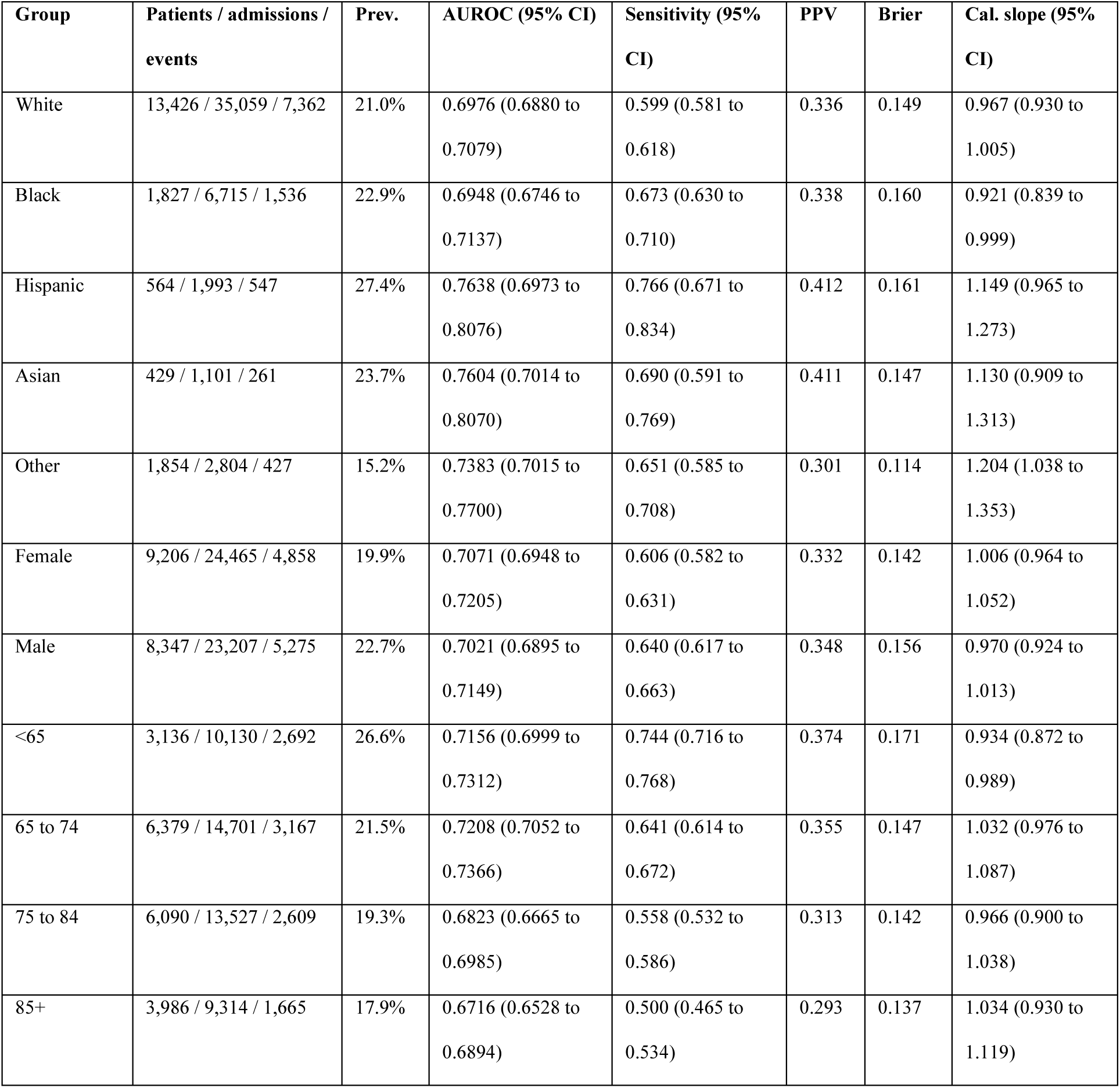
Subgroup performance on the test partition at the development-derived threshold (0.207). CIs are 95% patient-cluster bootstrap intervals; complete metrics appear in Supplementary Appendix 1. Hispanic: Hispanic or Latino; Other: other or unknown race (heterogeneous, not interpreted). PPV: positive predictive value; Prev.: prevalence; Cal.: calibration. Patients whose recorded race or age band changes between admissions appear in more than one row of a block.

**Figure 5.**
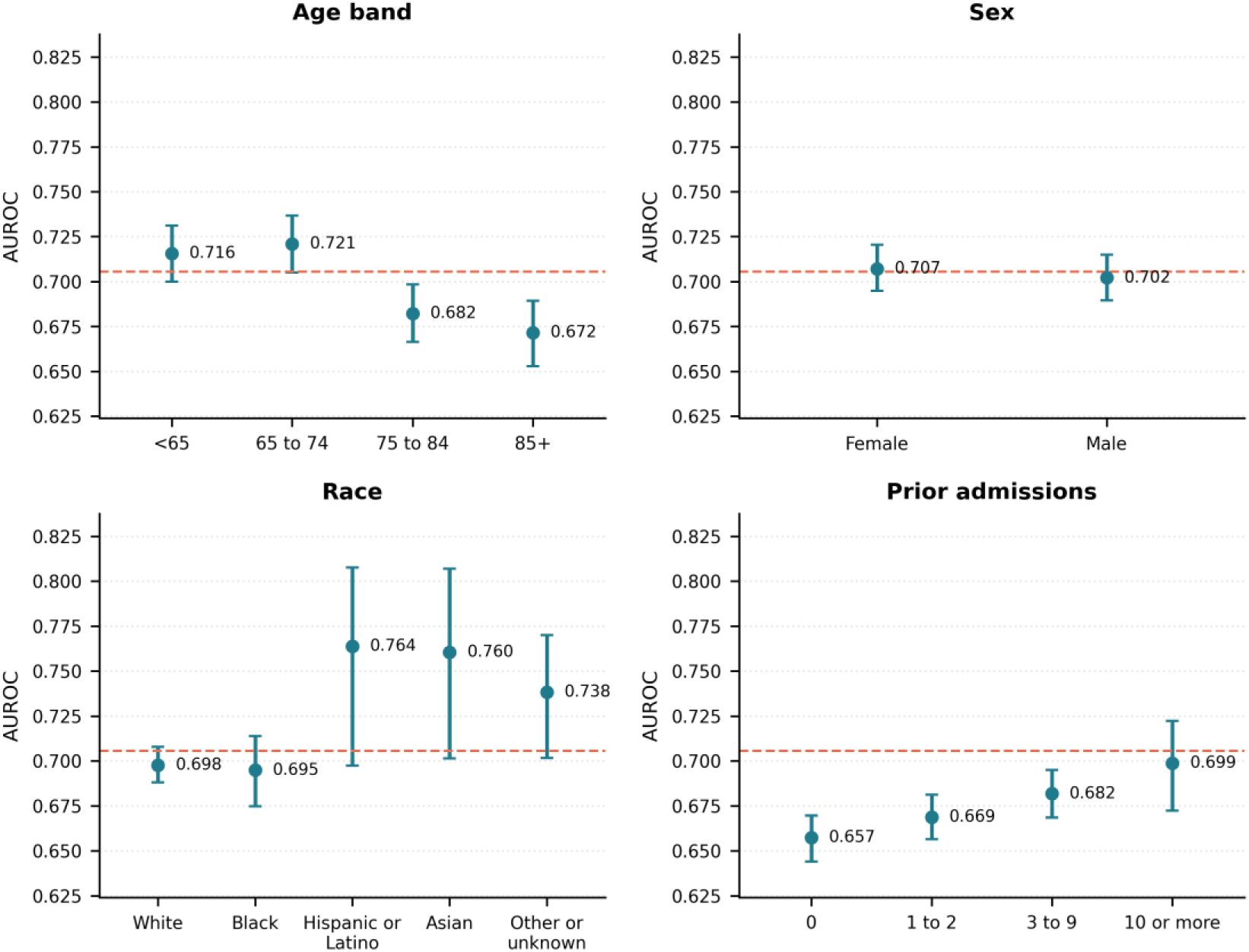
Test AUROC by age band, sex, race, and number of prior admissions, with 95% patient-cluster bootstrap CIs; the dashed line marks the overall test AUROC. Alt text: Four panels of AUROC point estimates with vertical 95% intervals by age band, sex, race, and number of prior admissions; a dashed horizontal line marks the overall test AUROC of 0.706. Intervals are widest for Hispanic, Asian, and other groups.

### Interpretability via SHAP

The leading predictors by mean absolute SHAP value were admission frequency by recency (0.227 on the log-odds scale), abnormal laboratory rate, laboratory results (log), prior admissions in the prior 6 months, and medication administrations (Figure 6). No single feature dominated: prior utilization, laboratory status at discharge, and the intensity of medication and laboratory activity during the stay jointly carried the signal.

**Figure 6.**
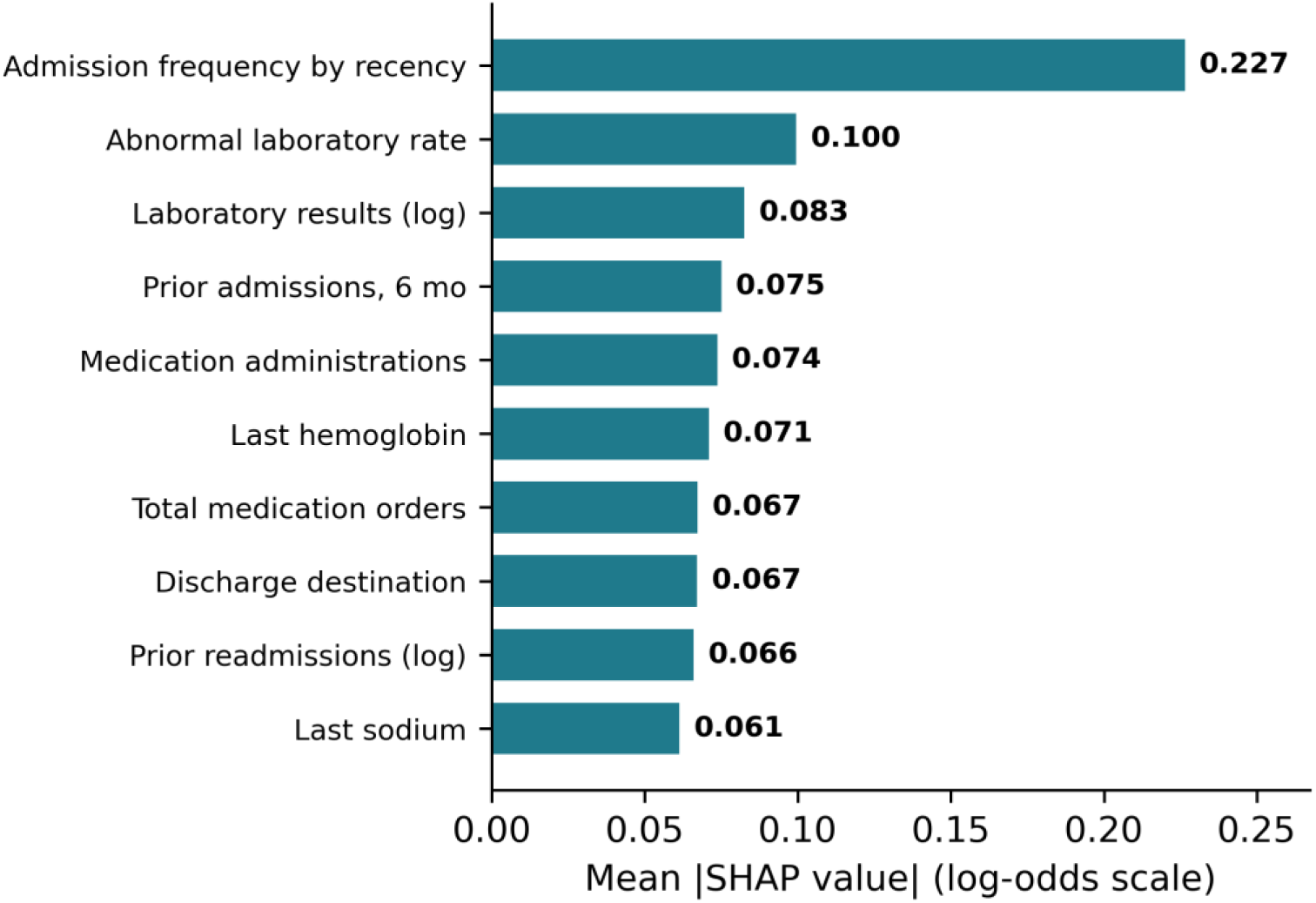
Top 10 predictors of the final 44-feature model by mean absolute SHAP value on the test partition (log-odds scale). Alt text: Horizontal bar chart of mean absolute SHAP values for the 10 leading features; the largest is admission frequency by recency at 0.227, followed by abnormal laboratory rate and laboratory results (log).

### Clinical Utility (Exploratory)

Decision curves showed net benefit above treat-none across thresholds 0.05 to 0.40 and above treat-all from 0.08 to 0.40 (Supplementary Appendix 4). Flagging the top 10% of admissions captured 25% of readmissions at a positive predictive value of 0.53. Patients who die without readmission are nonevents, so low risk is not clinical stability. Net benefit is not evidence that use would improve care.

## Discussion

### Principal Findings

In 236,906 Medicare-insured admissions, the most general finding (RQ5) is that a feature which passed a documentation-based audit and patient-grouped validation encoded later admissions; it inflated AUROC by about 0.07 and created an apparent advantage for first observed admissions, a rising calendar-period gradient, and a racial discrimination gap, all of which disappeared once it was removed. A separate outcome artifact, probable administrative continuations within an hour of discharge, accounted for most of the day-1 discrimination peak. The corrected 44-feature model reached an out-of-fold AUROC of 0.706, moderate discrimination, with near-ideal calibration on the exposed test partition; its contribution is methodological, not a deployable tool. First (RQ1, RQ3), risk was carried jointly by recency-weighted prior utilization, laboratory status at discharge, and in-hospital care intensity. Second (RQ2), on the test partition the model exceeded LACE and HOSPITAL, including versions refit to this cohort, by 0.04 to 0.10 AUROC; the scores target other outcomes, so the advantage is modest and not decisive. Third (RQ4), once probable administrative continuations were separated from readmissions, discrimination was lowest in the first week and rose later in the window; no difference by race or sex was detected, but specificity was lower for Black patients, and discrimination was lower in older patients and in first admissions.

### Lessons for Leakage Auditing

The leaked feature was documented as a prior 180-day window and passed the availability audit; patient-grouped validation could not expose it because the future information was present in every partition. Inexpensive checks caught it: testing history features among first observed admissions, flagging features constant within patients, and recomputing time-windowed features from raw timestamps. The first two are screens, not proof: constancy within patients also describes legitimate unit flags, and a first-admission test cannot detect every form of future information; recomputation from raw timestamps is the decisive step; comparing stored columns with raw tables also exposed median imputation fitted on all partitions, a quieter defect that leaks only the predictor distribution but contradicts a fold-local claim. These checks do not test whether index-stay features include events after discharge, which requires event-level timestamps. Similarly, 4.7% of outcome events were probable administrative continuations within an hour of discharge; inspecting the distribution of discharge-to-readmission gaps is a simple safeguard before any timing claim.

### Comparison With Prior Work

On MIMIC-IV, our out-of-fold AUROC of 0.706 is comparable to the structured-feature report (0.696) [32] and below notes-based (0.727) [31] and multimodal (0.791) [30] reports and full-EHR deep learning elsewhere (0.75 to 0.76) [14], which differ in cohort, outcome, and validation design. We cannot exclude similar leakage in published models that engineer history features.

### Clinical Interpretation (Hypotheses)

SHAP values describe the model’s reliance on a feature, not causal effects. Recency-weighted prior admissions are associated with return, possibly through unmet post-discharge needs or outpatient-access barriers, so the score should be paired with additional resources, never reduced care [5]. Discrimination is weakest for first admissions, where no history exists, and in the first post-discharge week, the window that transitional interventions target; early follow-up decisions should not rest on the score alone.

### Limitations

This is a single-center, retrospective, internally validated study whose case mix (73.5% of test admissions White) differs from national Medicare; no clinical-impact evidence exists. The outcome is within-system, so out-of-system readmissions are missed, and planned readmissions are included. The model family and hyperparameters were chosen during earlier development, which had evaluated models on the test partition, and were not re-selected within the outer folds, so the out-of-fold estimate is not fully independent of those choices. The test partition was used repeatedly in earlier development, and the leak itself was found only when we re-examined an earlier version’s features; the present estimates follow a principled correction rather than a fresh holdout, and residual indirect influence cannot be excluded. The validation partition participated in consensus selection and set the threshold. Earlier analysis versions used the globally imputed feature table; it was rebuilt before this version. The original code of the leaked feature, recovered in a final audit, confirms the mechanism the checks inferred. Order and transfer features were extracted with a discharge-time filter in the source pipeline and spot-checked, not fully recomputed, against raw tables. The primary outcome retains probable administrative continuations (4.7% of events). Social determinants are unavailable. Bootstrap intervals do not capture retraining or reselection variability.

## Conclusion

A documented history feature that encoded later admissions survived an availability audit and patient-grouped validation, inflated discrimination, and produced spurious subgroup and calendar-period findings; first-admission tests, within-patient constancy checks, and recomputation from raw timestamps exposed it and should accompany availability audits. After correction, a calibrated 44-feature model estimates 30-day within-system readmission in Medicare-insured adults with an out-of-fold AUROC of 0.706, modestly above reconstructed and refit bedside scores. External validation and prospective evaluation are required before clinical use.

## Data Availability

MIMIC-IV v3.1 is available from PhysioNet under a credentialed data use agreement and is not redistributed. The analysis code from the source feature table onward, the leakage tests, and the machine-readable results files from which every value in this manuscript is generated are available at https://github.com/thiagobandeira1/Medicare-30day-Readmission-MIMIC-IV and archived at Zenodo under concept DOI 10.5281/zenodo.21987701.

## Acknowledgments

This work began as the capstone project for the Master of Science in Data Science and Artificial Intelligence at Florida International University. We thank the MIT Laboratory for Computational Physiology and the PhysioNet team for maintaining MIMIC-IV. Generative AI tools (Anthropic Claude and OpenAI ChatGPT) were used for coding, writing, and review assistance; the authors take full responsibility for the content.

## Conflict of Interest Statement

None declared.

## Funding

This study received no external funding.

## Data Availability Statement

MIMIC-IV v3.1 is available from PhysioNet under a credentialed data use agreement [29,34] and is not redistributed. The analysis code from the source feature table onward, the leakage tests, and the machine-readable results files from which every value in this manuscript is generated (final_model_v13.json, the v8 to v13 audit files, and companions) are available at https://github.com/thiagobandeira1/Medicare-30day-Readmission-MIMIC-IV and archived at Zenodo under concept DOI 10.5281/zenodo.21987701; Zenodo versions under that DOI dated before October 2026 predate the corrections. A demonstration prototype in the companion repositories (https://github.com/thiagobandeira1/readmission-risk-api and https://github.com/thiagobandeira1/riskpath-clinician-companion) illustrates an earlier development model and is not the model reported here.

## Author Contributions

Conceptualization: TB; methodology: TB, AG, CP, AMM; software, formal analysis, and visualization: TB, AG; data curation: TB, AG; investigation and validation: TB, AG; writing (original draft): TB; writing (review and editing): AG, CP, AMM; supervision: CP, AMM. All authors approved the final version. (CRediT taxonomy.)

## Supplementary Material

Supplementary Material (single file): Supplementary Methods (cohort and outcome definitions, continuation band, partitions, leakage test, timestamp audit, missing-data handling, clinical-score reconstruction and refits, time-to-event and statistical detail), Supplementary Results (sensitivity and calendar-period analyses, payer scope, fixed-set comparison, robustness checks, tie rule, prior-utilization, race, and subgroup calibration analyses, stage attribution), Supplementary Tables S1 to S4 (calendar period, prior-utilization strata, definitions of the final features, and episode flow), and Supplementary Figures S1 to S3. Supplementary Appendix 1: predictor audit and selection workbook. Supplementary Appendix 2: LACE and HOSPITAL reconstruction. Supplementary Appendix 3: race consolidation. Supplementary Appendix 4: decision curves and development history. Supplementary Appendix 5: TRIPOD+AI checklist. Supplementary Appendix 6: PROBAST+AI self-assessment.

## Supplementary Material

Supplementary Appendices 1 to 6 are separate files listed in the main manuscript. All values below are generated from the released results files (results_v13 and the retained v8 to v10 audit files in results_v7).

### Supplementary Methods

#### S1. Cohort, Outcome, and Continuation Band

The outcome was the first subsequent hospitalization of the same patient in the same health system beginning more than 0 and up to 30 days (day 30 inclusive) after index discharge, of any cause and irrespective of how it ended or of the insurance recorded on it. The interval Δ is the exact difference between the next admission time and the index discharge time. Next admissions with −2<Δ≤0 days (n=650; none overlaps by more than 2 days) were treated as transfers. Admissions followed by death without readmission are nonevents in the binary frame. Because MIMIC-IV is single-center, the outcome is within-system.

Continuation band. Among all 50,793 outcome events, 2,368 (4.7%; 462 in the test partition) began within 1 hour of discharge, a rate of 2,368 per hour, against 44 per hour at 1 to 2 hours, 41 at 2 to 4 hours, 94 at 6 to 12 hours, and 116 at 1 to 3 days. Of these early records, 51.0% were internal transfers to or from psychiatry, 14.4% arrivals from skilled nursing, and 40.4% carried an emergency department registration that began before the index discharge; the index discharge destination was missing for 62.6%, hospice for 14.4%, and a psychiatric facility for 16.1%. By contrast, records beginning 1 to 12 hours after discharge arrived predominantly through the emergency department after a typical emergency stay, consistent with genuine early returns. These features make most early records probable administrative continuations, but they do not establish that each one is; and the 1-hour window was chosen after inspecting the data. The primary definition was therefore retained. The 1-hour sensitivity rule treats each continuation as part of the index stay. In the raw timestamps the median gap between the index discharge and the continuation was 1 minute (90th percentile 8 minutes) and the continuation lasted a median of 7.0 days (8.5% lasted beyond day 30), so timing the window from the first discharge would count hospital days as time at risk. The rule therefore merges every consecutive admission beginning within 1 hour of the previous discharge into one stay (176 stays had more than one link), starts the 30-day window at the discharge of the last link, excludes stays ending in death (285; 54 in test), as index in-hospital deaths are excluded from the cohort, and counts the next admission if it begins within that window. Features and predictions remain those of the first discharge. Of the 1,914 remaining stays, 330 (63 in test) were followed by a genuine return and remain events; simply relabeling every continuation as a nonevent would misclassify these returns (frozen-model AUROC 0.6946 under that cruder rule). Under the episode rule (9,676 test events, 20.5%), the frozen final model scores 0.6977 (95% CI 0.6880 to 0.7054) and a model refit to this outcome 0.7019 (95% CI 0.6922 to 0.7102); part of the frozen model’s primary AUROC therefore reflects the continuations.

**Table S4.**
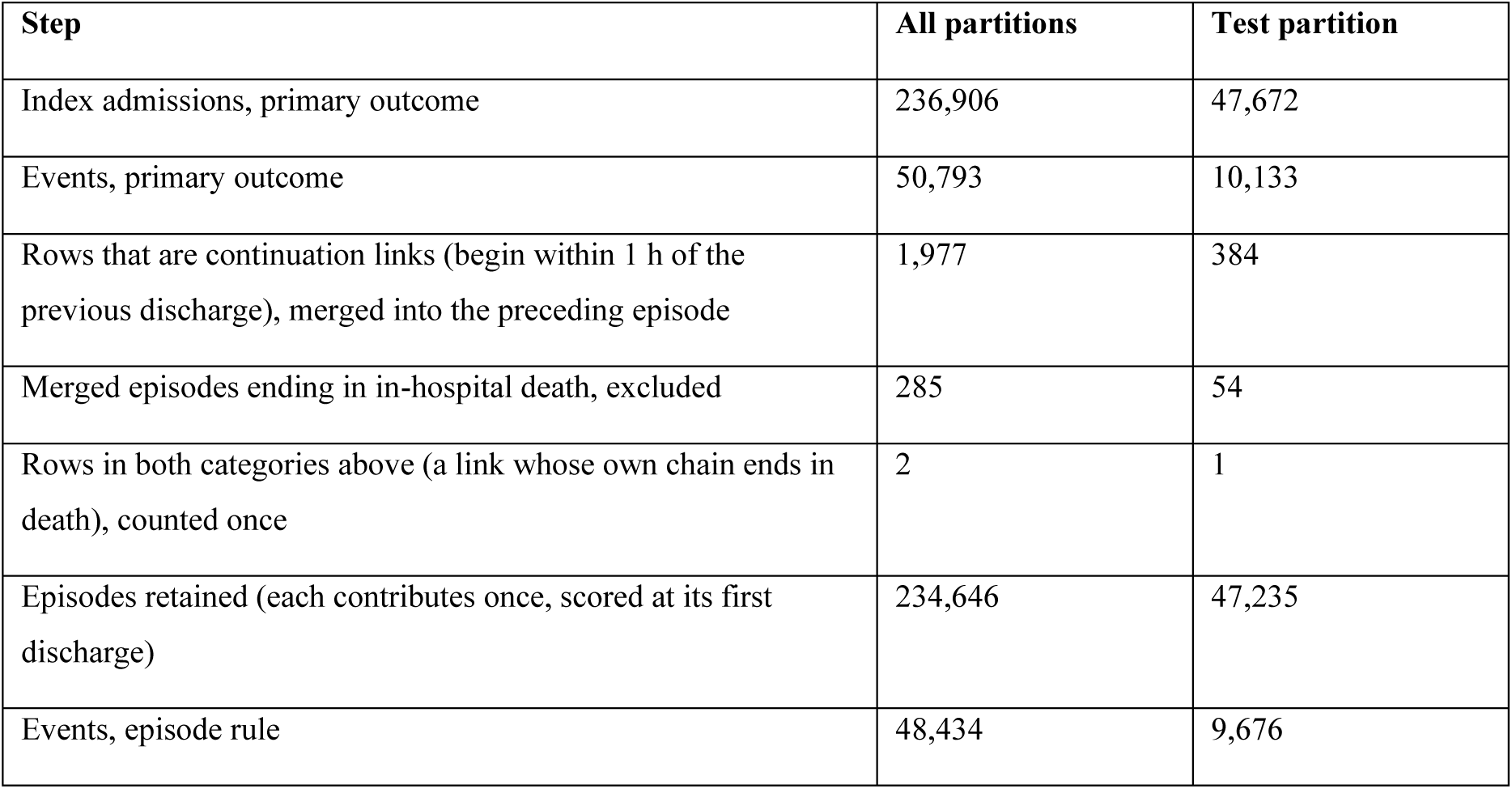
Episode flow under the 1-hour continuation sensitivity rule. Retained episodes equal index admissions minus merged links minus death exclusions plus the rows counted in both categories.

| **Step** | **All partitions** | **Test partition** |
|---|---|---|
| Index admissions, primary outcome | 236,906 | 47,672 |
| Events, primary outcome | 50,793 | 10,133 |
| Rows that are continuation links (begin within 1 h of the previous discharge), merged into the preceding episode | 1,977 | 384 |
| Merged episodes ending in in-hospital death, excluded | 285 | 54 |
| Rows in both categories above (a link whose own chain ends in death), counted once | 2 | 1 |
| Episodes retained (each contributes once, scored at its first discharge) | 234,646 | 47,235 |
| Events, episode rule | 48,434 | 9,676 |

#### S2. Partitions and Data Flow

Admissions were split 80/20 at the patient level (GroupShuffleSplit on subject_id); 10% of the development portion formed the validation partition (18,557 admissions, 7,042 patients, 3,995 events). The 5 outer selection folds span all development patients; within each outer-training fold, 3 grouped inner folds drive staged elimination with encodings refit from inner-training patients at every step; each fold’s selected model is refit on its outer-training fold and scored once on its outer-evaluation fold (primary estimate); the consensus set is formed from the 5 selections; and the final model is refit on the training partition, thresholded on the validation partition, and scored once on test. The validation partition therefore participated in consensus selection.

#### S3. Empirical Future-Information Tests and Timestamp Audit

Among 78,419 first observed index admissions (no earlier admission in MIMIC-IV), 51.5% had a later admission. For each of the 207 candidates we compared the share of rows at the feature’s modal value between patients without and with a later admission. For a feature built only from earlier encounters this test has a sharp prediction (a single fixed value for first observed admissions); for other features, such as index-stay measurements or age, differences are expected and the test is not informative. A prior-only feature should show no systematic difference; a feature fixed at one value for every patient without a later admission, but not for others, encodes the future. Only the 180-day length-of-stay trend met this criterion: it was nonzero for 44.2% of first observed admissions, every one of which had a later admission (0.0% of patients without one had a nonzero value; AUROC 0.929 for a later admission). Thirty-day readmission was 29.8% when the trend was nonzero and 3.1% when it was zero. The trend was also identical across all admissions of every one of the 45,154 cohort patients with more than one admission (100.0%) and zero for every patient with a single admission, so it was a per-patient summary of the whole longitudinal record. A final audit recovered the original capstone code, which confirms this: for each admission it took the length of stay minus that of the patient’s previous admission, averaged these differences over the patient’s whole record, and attached the mean to every admission (zero for patients with one admission), with no 180-day window. Applied to the source table, it reproduces the stored values for 100% of its 244,576 admissions. The trend and the two interaction terms built from it were removed. To test whether the trend concept itself carried signal, we rebuilt it causally: the least-squares slope of length of stay across the patient’s stays admitted within 180 days before, and discharged before, the index admission, followed by the index stay (0 when no such stay exists), with the same two interactions. The rebuilt trend was zero for all first observed admissions and, among the 40.6% of admissions with an earlier stay in the window, was unrelated to later admission (AUROC 0.513). Added to the final features (both sets refit in the same run) it changed development cross-validation AUROC from 0.7057 to 0.7056 and validation-partition AUROC from 0.7169 to 0.7172, so it was not added. The largest gap among the 44 final features was 0.039 (age).

Within-patient constancy. Among the 45,154 patients with more than one admission, the removed trend was constant within 100.0% of patients. Sex is constant by construction (recorded once per patient) and is a legitimate predictor; among the other final features the highest share was 81.4% for a binary unit flag. Constancy is a screen, not evidence of leakage; it is combined with the first-admission test and recomputation from raw timestamps.

Historical audit of the source table (v8; the discharge interval was later redefined in v13, below). Recomputation of history features from raw admission timestamps (all 244,576 cohort admissions, using only admissions before the index admission): prior admissions of any insurance 100.0%; admissions discharged in the prior 180 days 99.7%; mean whole-day length of stay of those admissions 99.8%; any prior 180-day admission with an emergency department registration 99.6%; days since the previous Medicare-insured discharge 99.99% (a fixed value whenever no earlier Medicare-insured admission exists, 100.0% agreement); and earlier Medicare-insured admissions in the prior 365 days followed by a 30-day readmission 100.0%. Definitions that would use later admissions matched worse (for prior readmissions, counting the index admission’s own outcome 67.3%, the whole record 54.1%). Several first-pass definitions did not match; each was resolved by a causal variant, and Supplementary Table S3 now gives the verified definitions.

Outpatient-record features (candidates, not in the final model) reproduced exactly as the last value dated on or before the admission time: body-mass index 100.0%, diastolic pressure 100.0%. Agreement with the last value dated on or before discharge was lower (86.5% and 97.2%) because 11.0% and 0.4% of admissions had a value charted during the stay, which the features correctly exclude.

Index-stay features (random sample of 3,000 admissions): numeric laboratory results per admission reproduced 100.0%, the abnormal laboratory rate 100.0%, distinct laboratory test types 94.9%, and distinct order types 99.8% (spot check made before the cutoff rebuild described below). The feature stored as orders_last_6h equals late order rate multiplied by late orders (100.0%), not orders in the final 6 hours as its name suggests. Rows recorded under an admission but timestamped after discharge were rare (0.02% of laboratory and 0.06% of order rows in a sample of 20,000 admissions; median 1.8 hours after discharge for orders) and unrelated to the outcome (AUROC 0.50 and 0.50). Laboratory value features (last hemoglobin, albumin, sodium, and white cell count; maximum bilirubin) were rebuilt for every admission from the raw laboratory table, keeping a result only if both its collection time and its storage time (storetime, when the result became available) fell on or before discharge; 6,205 of 14,905,537 results (0.04%) collected before discharge were stored afterwards and are excluded. The laboratory counts (results, abnormal rate, test types, critical results) and the prescription count, which the source pipeline had taken from every row of the admission, were rebuilt for every admission under the same cutoffs; 4.0% of laboratory result counts and 22.8% of medication counts changed (mean results 167.0 to 166.7; Supplementary Appendix 1, Cutoff_Rebuild). Two features that were exact copies of others under different names (orders_first_24h, identical to orders per day; nonenglish_x_prior_admits, identical to prior admissions in 6 months) were removed from the eligible pool.

Documentation time. MIMIC-IV records when an event occurred (charttime, starttime) and when it was documented (storetime for chart and medication-administration records; pharmacy entertime for prescriptions). The source pipeline filtered these tables on event time only. Every chart, medication-administration, and prescription feature was therefore rebuilt from the raw tables requiring both times to fall on or before discharge. Of records already filtered by event time, 3,662 of 21,614,955 medication-administration records (0.017%), 16,621 of 11,589,196 chart records for the selected items (0.143%), and 161 of 10,469,390 prescriptions (0.002%) were documented after discharge and are now excluded. Before the cutoff the recomputed final-model features reproduced the stored values for 99.3% to 100% of admissions, confirming the definitions (two eligible candidates outside the final set, incomplete-dose count and rate, reproduced 98.4% because the source parsed the incomplete-dose field differently); the cutoff changed 1.22% of values at most (per feature in Supplementary Appendix 1, Cutoff_Rebuild, which also records the v12 count rebuild). The discharge interval was also recomputed from any earlier admission (4.2% of values changed). Order and transfer features were extracted with a discharge-time filter in the source pipeline (ordertime, transfertime, or intime on or before discharge) and were spot-checked, not fully recomputed, against raw tables; the provider order entry table carries no separate documentation time.

#### S4. Missing-Data Handling

The source feature table, inherited from the capstone pipeline, had no missing values: continuous measurements that were never taken had been imputed with the median computed over all 244,576 admissions, including the test partition, and some had been zero-filled. Comparing stored columns with raw tables identified 16 median-imputed features (last creatinine, maximum creatinine, last sodium, last potassium, last white cell count, last glucose, maximum glucose, last bnp, maximum lactate, maximum inr, last bicarbonate, last magnesium, last phosphate, heart rate variability, systolic bp variability, minimum gcs) and 5 zero-filled features (creatinine change, last bun, last hemoglobin, last albumin, maximum bilirubin). All were rebuilt from raw MIMIC-IV tables with missing values preserved; true missingness ranged from 13.9% to 93.5%, and where a measurement existed the old and new values agreed for 99.8% or more of admissions (the remainder reflect the storage-time rule). Intensive care hours and the longest unit stay are zero when no intensive care unit stay occurred. Counts are zero when no event occurred. Time since last discharge is the interval from the previous discharge of any insurance and is missing only for first observed admissions (33.4% of admissions; the source table had used the previous Medicare-insured discharge and the global median, 80.22 days, when none existed); recency-weighted frequency is zero for first observed admissions. Interaction terms are missing when a component is missing. Percentages in this paragraph that come from the rebuild report use all 244,576 Medicare admissions as denominator; the audit workbook reports missingness on the analysis cohort of 236,906 admissions, which is why its values differ slightly. The tree learner handles missing values natively. The logistic comparator used fold-local median imputation with standardization. The global imputation leaked only the distribution of predictors across partitions, not outcomes; the complete pipeline was rerun on the rebuilt table.

#### S5. Clinical-Score Reconstruction and Refits

Adapted LACE (van Walraven et al.): length-of-stay points by the original bands; 3 acuity points unless elective; Charlson comorbidity points from index ICD diagnoses; and prior 180-day emergency department visits capped at 4, counting only visits that led to admission (emergency department visits without admission are not in the MIMIC-IV core tables, so the score is adapted and may understate the E component). HOSPITAL (Donze et al.): hemoglobin below 12 g/dL, oncology service, sodium below 135 mmol/L, any ICD-coded procedure, nonelective admission, prior admissions in 12 months, and length of stay of 5 days or more. Recomputed scores matched the stored test scores for 99.9% (LACE) and 100.0% (HOSPITAL) of admissions. Refits used logistic regression on indicator-coded components, fitted on the training partition: LACE all components AUROC 0.6400, without the coded comorbidity item 0.6381; HOSPITAL all components 0.6592, without the coded procedure item 0.6573; both scores’ discharge-available components together 0.6647. Uplifts of the final model over these refits were 0.0656, 0.0674, 0.0463, 0.0483, and 0.0409 (all 95% CIs excluding zero; descriptive, no P values computed; Supplementary Appendix 1). On the development partitions, out of fold, adapted LACE scored 0.6055 and HOSPITAL 0.6424 against 0.7057 for the selection procedure (paired uplifts 0.1002, 95% CI 0.0961 to 0.1041, and 0.0634, 95% CI 0.0607 to 0.0662). Top-decile event capture out of fold was 25% for the procedure, 16% for LACE, and 20% for HOSPITAL.

#### S6. Time-to-Event Detail

Events were readmission (event 1), death before readmission within 30 days (event 2; registry deaths beyond 1 year after last discharge are censored by deidentification), and administrative censoring at day 30. The AFT model used early stopping on the validation partition (best iteration 398). Daily cumulative/dynamic AUROC keeps deaths before day t among the controls (Blanche definition 2); with no censoring before day 30, the estimate is the empirical AUROC. Treating deaths as censored at death with inverse probability weighting gave nearly identical values (day 1 0.8364 vs 0.8363). Landmark models were trained on training-partition patients still at risk when each stage opens and evaluated on the test partition: primary definition 0.6867 (4,000 events among 47,672), 0.6883 (2,545 events among 43,236), 0.7103 (3,572 events among 40,375); 1-hour episode rule 0.6619, 0.6872, 0.7110 for days 1 to 7, 8 to 14, and 15 to 30.

#### S7. Bootstrap and Subgroup Detail

Resample counts: 1,000 for the test panel; 800 for subgroup and prior-utilization CIs; 500 for the primary out-of-fold estimate, model comparisons, and daily AUROC bands; 5,000 for contrasts with P values. Intervals resample patients and their stored predictions; selection and fitting were not repeated within draws. The White vs Black contrasts resample the union of the two patient sets jointly, so a patient recorded in both groups receives the same multiplicity in both. Race strings were consolidated by substring (WHITE to White; BLACK to Black; HISPANIC or LATINO to Hispanic or Latino; ASIAN to Asian; all others, declined, or unknown to Other or unknown). Prior admissions count every earlier admission of the patient in MIMIC-IV regardless of insurance.

#### S8. Demonstration Prototype

The public demonstration prototype is documented as a research prototype, not a medical device. It runs an earlier development model, not the model reported here.

### Supplementary Results

#### S9. Sensitivity and Calendar-Period Analyses

(1) Counting the transfer band as readmissions raised test events from 10,133 to 10,273; refit AUROC 0.7101. (2) Excluding next admissions typed ELECTIVE (9,852 events, 20.7%) gave 0.7000. (3) Nonelective index admissions only (43,012): 0.7073. (4) Excluding 6,658 test admissions (14.0%) whose latest possible year reached 2022: 0.7059. (1) and (2) are models refit to the revised outcome; (3) and (4) evaluate the final model on subsets. (5) Treating returns within 1 hour as part of the index stay (episode rule, Table S4; 9,676 test events, 20.5%): refit 0.7019, final model 0.6977 (S1). (6) Calendar-period bands appear in Table S1.

**Table S1.** Test discrimination by patient-level anchor_year_group band. eMAR coverage is the share of admissions with at least one medication-administration record. Bands are descriptive.

| **Calendar period** | **Test admissions** | **AUROC** | **First admissions** | **AUROC, first admissions** | **eMAR coverage** |
|---|---|---|---|---|---|
| 2008 to 2010 | 22,268 | 0.7063 | 21.2% | 0.6535 | 31.9% |
| 2011 to 2013 | 9,806 | 0.6939 | 31.9% | 0.6694 | 46.1% |
| 2014 to 2016 | 7,062 | 0.6931 | 41.3% | 0.6446 | 89.9% |
| 2017 to 2019 | 5,283 | 0.6952 | 52.0% | 0.6463 | 98.9% |
| 2020 to 2022 | 3,253 | 0.6963 | 65.1% | 0.6705 | 98.4% |

Discrimination was stable across calendar periods despite large differences in medication-administration data coverage. Before the leak was removed, test AUROC had risen from 0.744 to 0.856 across these bands, a gradient that disappeared with the leaked features.

#### S10. Payer Scope of Outcome Ascertainment

743 next admissions within 30 days carried non-Medicare insurance; 741 were counted as readmissions, the remainder falling in the transfer band.

#### S11. Fixed-Set Comparison and Robustness Checks

With both predictor sets fixed, grouped cross-validation on identical development folds gave AUROC 0.7057 for the 44-feature consensus set and 0.7065 for the 137-feature pool (difference −0.0008, 95% CI −0.0017 to 0.0001). Consensus cross-validation folds were 0.7066, 0.7033, 0.7040, 0.7056, 0.7090.

Post hoc robustness checks on the validation partition (not used to change the model): with the final features fitted on the training partition, the fixed hyperparameters gave AUROC 0.7177; 14 alternative settings (maximum depth 3 to 8, learning rate 0.03 to 0.1 with matched tree counts) gave 0.7108 to 0.7178; five further seeds gave 0.7164 to 0.7174; and training-set AUROC was 0.7585. Using 10%, 50%, and 100% of training patients gave 0.6799, 0.7112, and 0.7177. A model restricted to the 6 prior-utilization features of the final set reached 0.6441 on validation and 0.6448 on test (one additional test evaluation; the full model’s advantage on test was 0.0608, 95% CI 0.0553 to 0.0660; descriptive).

#### S12. Same-Date Readmission and Death

Registry death dates are day-granular; same-date readmission and death were resolved death-first in the primary analysis. Resolving all 66 ties (16 in test) readmission-first changed Harrell C from 0.6757 to 0.6761 and the 30-day readmission cumulative incidence from 21.2% to 21.3%.

#### S13. Prior Utilization, Race, and Subgroup Calibration

**Table S2.** Test performance by number of prior admissions of any insurance.

| **Prior admissions** | **Admissions (share)** | **Prevalence** | **AUROC (95% CI)** | **Share of top-decile flags** |
|---|---|---|---|---|
| 0 | 15,639 (32.8%) | 14.6% | 0.6573 (0.6439 to 0.6695) | 6.1% |
| 1 to 2 | 13,778 (28.9%) | 18.2% | 0.6686 (0.6565 to 0.6811) | 11.9% |
| 3 to 9 | 12,780 (26.8%) | 25.4% | 0.6817 (0.6685 to 0.6948) | 35.6% |
| 10 or more | 5,475 (11.5%) | 38.2% | 0.6987 (0.6722 to 0.7222) | 46.4% |

White minus Black AUROC differences within strata were −0.009 (0), −0.001 (1 to 2), −0.000 (3 to 9), +0.041 (10 or more). Standardized to the White utilization mix, the difference was 0.001 (95% CI −0.017 to 0.022); crude (observed, unstandardized) 0.0029. Median prior admissions were 1 for White and 3 for Black patients. Every stratum has an AUROC below the overall 0.7056 because part of the model’s discrimination comes from separating patients between utilization levels, whose readmission rates differ sharply.

Calibration-in-the-large by subgroup (observed to expected ratio, 95% patient-cluster bootstrap CI, 800 resamples): Asian 1.03 (0.93 to 1.12); Black 1.00 (0.96 to 1.04); Hispanic or Latino 1.05 (0.94 to 1.13); other or unknown 0.78 (0.69 to 0.85); White 1.00 (0.98 to 1.02); by age band <65 1.00; 65 to 74 1.00; 75 to 84 0.98; 85+ 0.99. On average calibration only the heterogeneous other or unknown group was miscalibrated (risk overestimated); the Black calibration-slope interval excludes 1 (Table 4). Specificity at the shared threshold was 0.685 for White and 0.610 for Black patients (difference 0.075, 95% CI 0.048 to 0.102; 5,000 joint draws; descriptive). 6 of 15,247 patients carry both race labels across admissions and appear in both groups; the contrasts therefore resample the union of patients jointly (point estimates are observed differences; bootstrap means appear in Supplementary Appendix 1). A nonsignificant AUROC difference does not establish equivalent performance; the White minus Black AUROC interval in the main text spans differences from −0.018 to 0.024 (observed 0.0029, bootstrap mean 0.0031, 5,000 joint draws).

#### S14. Stage-Specific Attribution (Exploratory)

Across days 1 to 7, 8 to 14, and 15 to 30, the prior-utilization share of attribution rose from 22.0% to 31.3%, while medication and order activity fell from 30.1% to 24.8% (Supplementary Figure S2, panel A). Features weighted more for early returns: late orders (+2.7 percentage points); discharge destination (+2.5 percentage points); last white cell count (+2.2 percentage points); prior readmissions (log) (+2.1 percentage points). Features weighted more for late returns: admission frequency by recency (−4.6); abnormal laboratory rate (−3.3); mean prior stay, 6 mo (−2.7); discharge medications (−2.6); laboratory results (log) (−2.3); prior admissions, 6 mo (−2.1). The 10 largest differentials were selected before interval inspection, so the intervals are descriptive. Stage models use the primary outcome definition.

#### Supplementary Table S3. Definitions of the Final Features

**Table S3.** The final features. Verification: share of admissions for which the stored value reproduces from its component features, from raw MIMIC-IV admission timestamps (all cohort admissions, earlier admissions only), or from raw event tables (random sample of 3,000 admissions); Documented: definition taken from the feature documentation and not recomputed.

| **Feature** | **Definition** | **Verification** |
|---|---|---|
| Abnormal laboratory rate | Laboratory rows flagged abnormal divided by numeric results, both charted and stored on or before discharge | Raw table, all admissions, rebuilt with discharge cutoff (v12) |
| Admission frequency by recency | Prior admissions divided by (days since last discharge + 1) | Components, 100% (v13 interval) |
| Admission source | MIMIC admission source; target-encoded fold-locally | Documented |
| Admission type | MIMIC admission type; target-encoded fold-locally | Documented |
| Age | Age in years at admission | Documented |
| Albumin by stay length | Last albumin multiplied by length of stay | Components, 100% |
| Antibiotic use | Any antibiotic administered | Raw table, all admissions, documentation-time cutoff (v13; 100.0% agreement before cutoff) |
| Consults per day | Consult orders per day of stay | Documented |
| Days since last discharge | Days from the previous discharge of any insurance to the index admission; missing for first observed admissions | Raw timestamps, 100% (v13 definition recomputed for every admission) |
| Days since last discharge (log) | Days since last discharge (log) | Components, 100% (v13 interval) |
| Discharge destination | Recorded discharge destination; target-encoded fold-locally | Documented |
| Discharge medications | Medications on the discharge list | Raw table, all admissions, documentation-time cutoff (v13; 99.9% agreement before cutoff) |
| Distinct drugs | Distinct drugs administered | Raw table, all admissions, documentation-time cutoff (v13; 99.3% agreement before cutoff) |
| Heart rate variability | Heart rate variability | Raw table, all admissions, documentation-time cutoff (v13; 100.0% agreement before cutoff) |
| IV administration rate | IV administration rate | Raw table, all admissions, documentation-time cutoff (v13; 100.0% agreement before cutoff) |
| Laboratory orders | Laboratory orders placed | Documented |
| Laboratory results | Numeric laboratory results charted and stored on or before discharge | Raw table, all admissions, rebuilt with discharge cutoff (v12) |
| Laboratory results (log) | log(1 + laboratory results) | Raw table, all admissions, rebuilt with discharge cutoff (v12) |
| Last albumin | Last albumin stored on or before discharge (g/dL); missing if none | Raw table, all admissions, stored before discharge (99.9% agreement) |
| Last hemoglobin | Last hemoglobin stored on or before discharge (g/dL); missing if none | Raw table, all admissions, stored before discharge (99.9% agreement) |
| Last sodium | Last sodium stored on or before discharge (mmol/L); missing if none | Raw table, all admissions, stored before discharge (99.8% agreement) |
| Last white cell count | Last white cell count stored on or before discharge; missing if none | Raw table, all admissions, stored before discharge (99.9% agreement) |
| Late order burden | Composite late-order burden | Documented |
| Late order rate | Late orders as a share of all orders | Documented |
| Late order rate by late orders | Late order rate multiplied by late orders (source column orders_last_6h) | Components, 100% |
| Late orders | Orders placed late in the stay | Documented |
| Length of stay | Length of stay in whole days (discharge minus admission, floored) | Documented |
| Length of stay (log) | log(1 + length of stay) | Components, 100% |
| Maximum bilirubin | Maximum total bilirubin stored on or before discharge; missing if none | Raw table, all admissions, stored before discharge (100.0% agreement) |
| Mean prior stay, 6 mo | Mean whole-day length of stay of earlier admissions discharged in the prior 180 days (0 if none) | Raw timestamps, 99.8% |
| Medication administrations | Medication administration (eMAR) records | Raw table, all admissions, documentation-time cutoff (v13; 100.0% agreement before cutoff) |
| Medication routes | Distinct medication administration routes | Raw table, all admissions, documentation-time cutoff (v13; 100.0% agreement before cutoff) |
| Midday discharge | Discharge between 10:00 and 14:59 (source column discharge_surge) | Documented |
| Minimum GCS | Minimum Glasgow Coma Scale charted during the stay; missing if never charted | Raw table, all admissions (100% agreement) |
| Order types | Distinct provider order types with ordertime on or before discharge (source extraction filter) | Raw table, sample, 99.8% |
| Orders per day | Provider orders per day of stay | Documented |
| Prior admissions, 6 mo | Earlier admissions (any insurance) discharged in the 180 days before the index admission | Raw timestamps, 99.7% |
| Prior admissions, 6 mo (squared) | Square of prior admissions in 6 months | Components, 100% |
| Prior admissions, all | Earlier admissions of the patient (any insurance) | Raw timestamps, 100% |
| Prior readmissions (log) | log(1 + prior readmissions) | Components, 100% |
| Provider fragmentation by stay length | Length of stay multiplied by distinct providers | Components, 100% |
| Stay length per prior admission | Length of stay divided by (prior admissions + 1) | Components, 100% |
| Step-down unit | Any step-down unit stay | Documented |
| Total medication orders | Prescriptions started on or before discharge and entered into the pharmacy system on or before discharge | Raw table, all admissions, documentation-time cutoff (v13; 100.0% agreement before cutoff) |

### Supplementary Figures

**Supplementary Figure S1.**
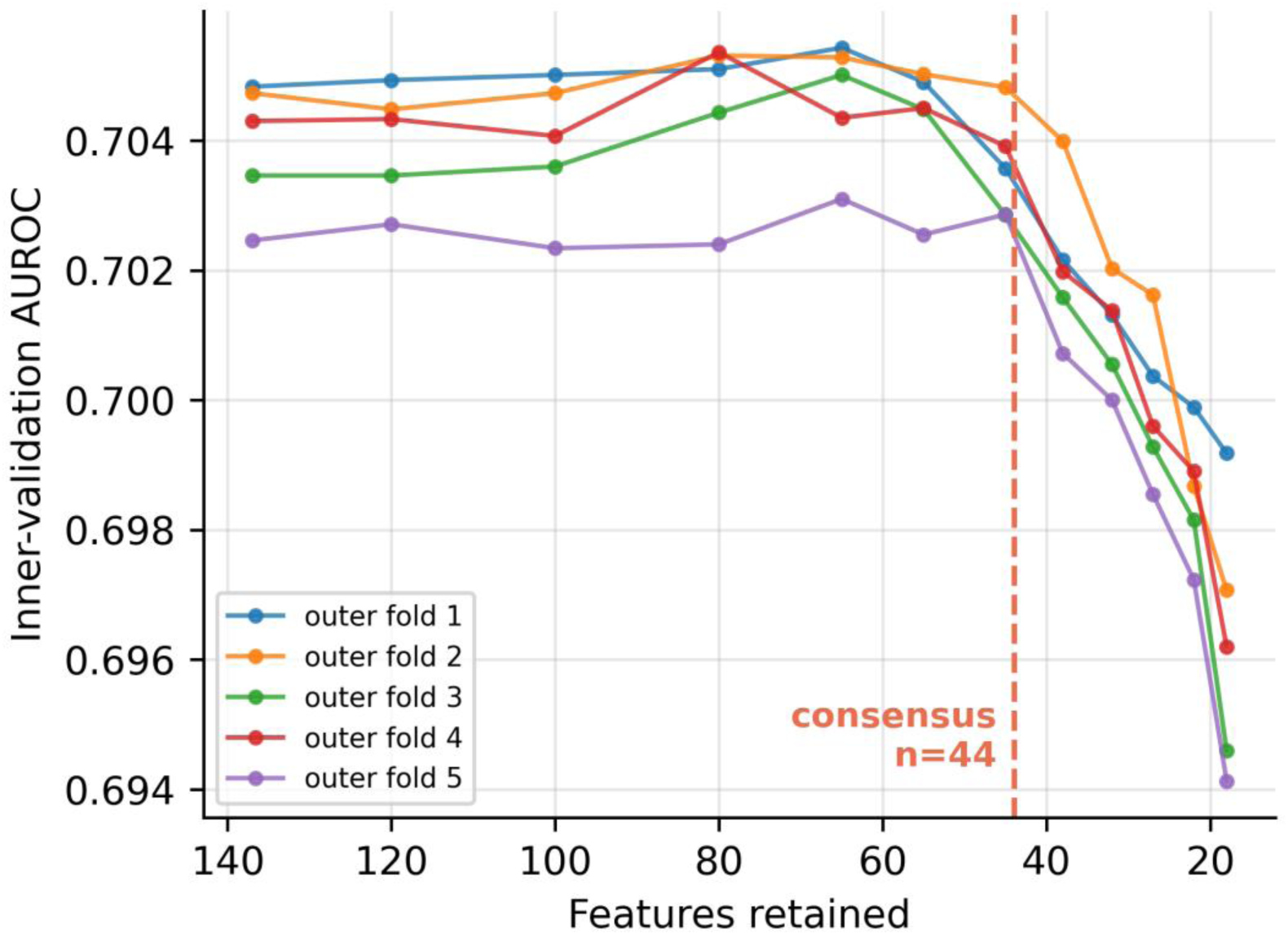
Staged recursive feature elimination on development data: mean inner-validation AUROC versus features retained, per outer fold; the dashed line marks the 44-feature consensus.

**Supplementary Figure S2.**
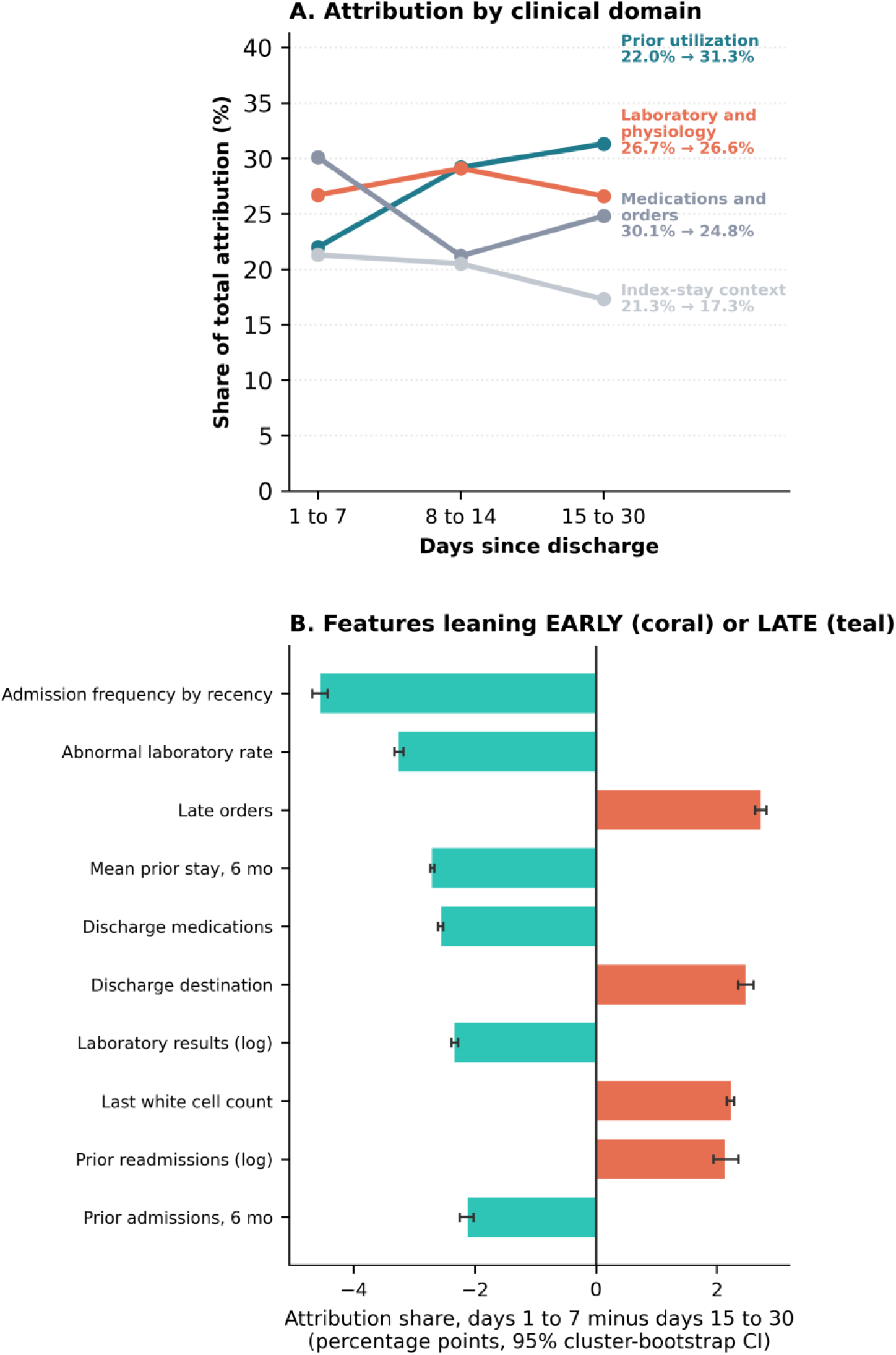
Attribution across post-discharge windows. (A) Share of total attribution by clinical domain. (B) Features whose attribution share differs most between days 1 to 7 and days 15 to 30; positive values indicate greater early importance; bars are 95% cluster-bootstrap CIs.

**Supplementary Figure S3.**
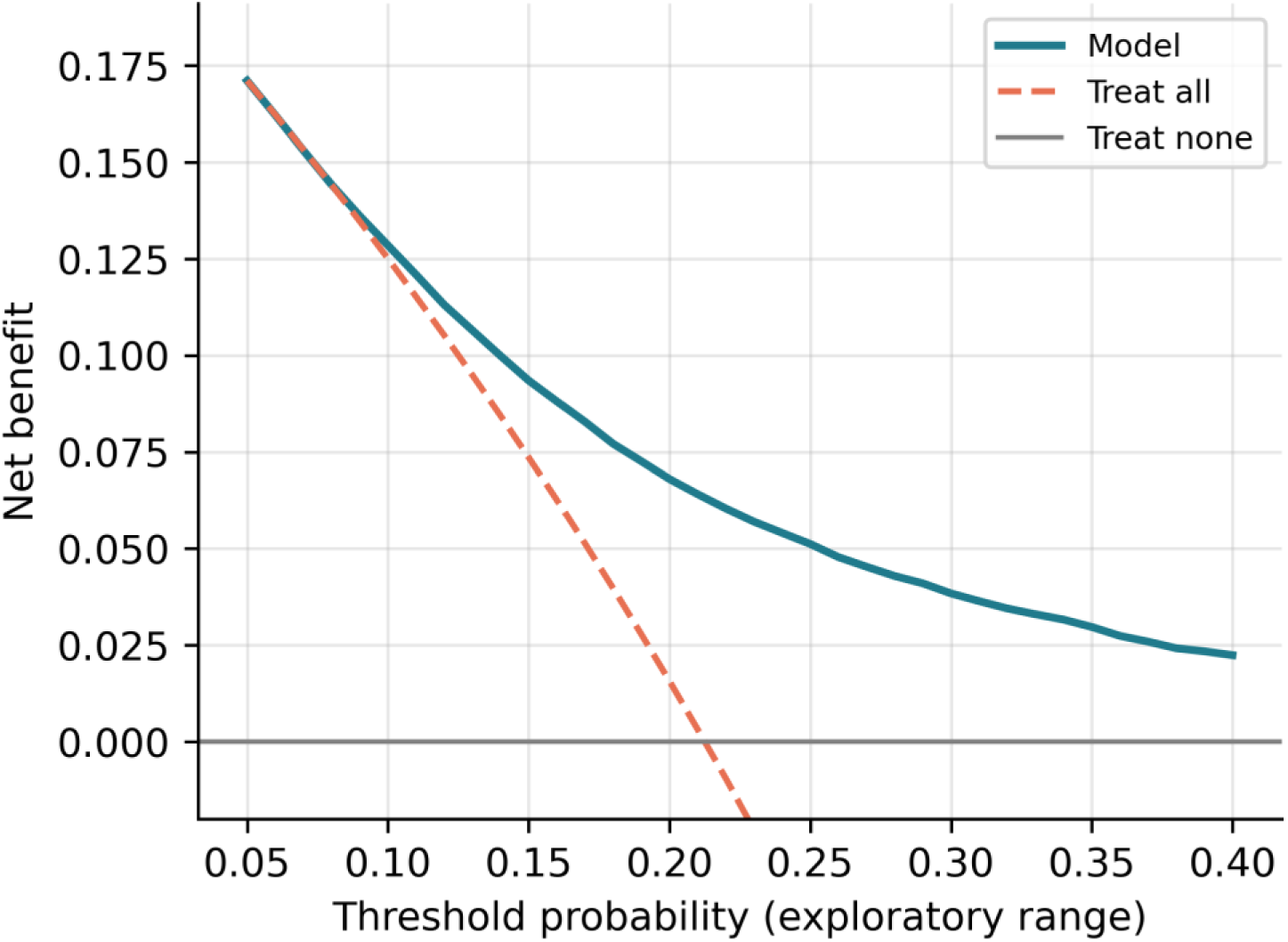
Decision curves for the final model on the test partition (exploratory threshold range).

## Notes

### Competing Interest Statement

The authors have declared no competing interest.

### Author Declarations

The Institutional Review Board of Beth Israel Deaconess Medical Center granted a waiver of informed consent for the MIMIC-IV database and approved its sharing. This study is a secondary analysis of the deidentified MIMIC-IV v3.1 data, accessed under the PhysioNet Credentialed Health Data Use Agreement; it did not require institutional review board review at Florida International University.

